# Use of penalized basis splines in estimation of characteristics of seasonal and sporadic infectious disease outbreaks

**DOI:** 10.1101/2020.07.14.20138180

**Authors:** Ben Artin, Daniel M. Weinberger, Virginia E. Pitzer, Joshua L. Warren

## Abstract

There is often a need to estimate the characteristics of epidemics or seasonality from infectious disease data. For instance, accurately estimating the start and end date of respiratory syncytial virus (RSV) epidemics can be used to optimize the initiation of prophylactic medication. These characteristics can sometimes be estimated directly from disease incidence data; more often, widely-used methods for describing these characteristics begin with a regression model fit to a time series of disease incidence. The fitted model is then used to calculate the quantities of interest. Calculation of these quantities typically involves combining multiple estimated parameters from the fitted model, and consequently only point estimates (rather than measures of uncertainty) can be made in a straightforward way. Motivated by attempts to estimate the optimal timing of prophylaxis for RSV, we developed a general method for obtaining confidence intervals for characteristics of seasonal and sporadic infectious disease outbreaks. To do this, we use multivariate sampling of a generalized additive model with penalized basis splines. Our approach provides robust confidence intervals regardless of the complexity of the calculations of the outcome measures, and it generalizes to other systems (including outbreaks of other infectious diseases). Here we present our general approach, its application to RSV, and an R package that provides a convenient interface for conducting and validating this type of analysis in other areas.

**Author summary:** Prevention and treatment of seasonal infections use numerous resources, such as pharmaceuticals, laboratory equipment, and clinical and non-clinical staff. Optimizing the use of these resources usually requires forecasting the timing of infectious disease seasons. In our research of respiratory syncytial virus (a seasonal respiratory infection that is a significant cause of infant hospitalizations and mortality world-wide) we used splines (a type of mathematical curves with convenient and well-understood statistical properties) to develop a new approach to obtain interval estimates of temporal characteristics of seasonal epidemics, such as the beginning and the end dates. We also developed an R package to facilitate use or our methods in other research. Here we present our general approach and outline its applications to RSV.

## Introduction

Analyses of infectious disease dynamics often make inferences based on timing for various quantities of clinical and public health interest; for example, the start and the end of seasonal outbreaks, duration of outbreaks, and/or optimal timing of prophylaxis or treatment. Some aspects of timing can be estimated ad-hoc directly from disease incidence data; more typically, a regression model (for example, a generalized linear model (GLM) or a generalized additive model (GAM)) is fitted to a time series of observed disease counts, and the calculations of the quantities of interest are made based on combinations of parameters estimated from the fitted model.

Calculating the uncertainty of estimates from these types of GAM/GLM-based methods presents a challenge under frequentist approaches. These challenges are exemplified by analyses of the dynamics of respiratory syncytial virus (RSV) — a respiratory infection that has annual or biennial seasonal pattern and is a major infectious cause of infant morbidity and mortality globally. [1, 2] In some countries, high-risk infants are administered antibody prophylaxis throughout the period of high RSV incidence. The therapy is expensive, and the duration of antibody protection is short-lived, [3, 4] so administration of prophylaxis needs to be timed with the RSV season to provide optimal protection. [5] Therefore, estimating the timing of initiation of the RSV season in different locations is important.

Frequentist model fitting procedures provide measures of uncertainty for the individual model parameters. However, calculating the quantities of interest from those model parameters often requires differentiation or integration with respect to time (for example, to identify outbreak transition points or to determine overall disease burden), which can complicate the process of deriving large sample standard errors. As a result, these tools in RSV research, such as those evaluated in [6], typically ignore uncertainty and focus solely on estimation. When they do consider uncertainty estimation, these methods in infectious disease modeling are often either complex to use (for example, block bootstrapping), computationally intensive, or both. Parameter resampling is an approach that offers uncertainty estimation, balances ease of use with computational efficiency, and has been previously applied in infectious disease modeling. [7, 8]

In this study, our goal was to generalize prior work on parameter resampling to develop a general framework of retrospective analysis of infectious diseases that allows estimation of uncertainty intervals for time-based outbreak characteristics (such as outbreak onset time) while being flexible as to the precise definition of the characteristics of interest. To that end, we will show how functional data analysis using GAMs with penalized splines can be used to fit smooth curves to infectious disease incidence data, and how the fitted model can then be used to calculate uncertainty intervals for characteristics of the epidemic, such as the start and end date of seasonal epidemics.

## Methods

We developed this method in the context of our RSV research, in which the principal questions were:

- Is the period during which RSV prophylaxis is administered well-matched to the timing of RSV seasons?
- Could the prophylaxis period be adjusted so that more of the prophylaxis-eligible RSV cases fall within the prophylaxis period?

To answer these questions, we defined the following outcome measures:

- Season onset and offset, defined as the points in time when the cumulative incidence of RSV rises above 2.5% and 97.5% of the total incidence of RSV for the entire season.
- Preventable fraction of a prophylaxis regimen, defined as the fraction of all prophylaxis-naive RSV cases occurring during the period when the prophylaxis regimen provides protection from RSV.

Our approach is to fit a family of smooth curves through the observed disease incidence time series, and to infer time-domain features of the outbreak from these curves using parameter resampling. The fitting curves we use are penalized basis splines (P-splines), which have several desirable qualities, including computational compactness and efficiency and well-understood statistical behavior. Most notably for our application, P-spline parameters estimated by GAM are asymptotically normally distributed, provided that the underlying phenomenon is well-approximated by a piecewise-polynomial function. [9] Additionally, we developed a simulation study to validate this interval estimation method for our outcome measures.

### Outcome estimation

By way of example, consider the (hypothetical) RSV season shown in Fig 1.

**Fig 1.**
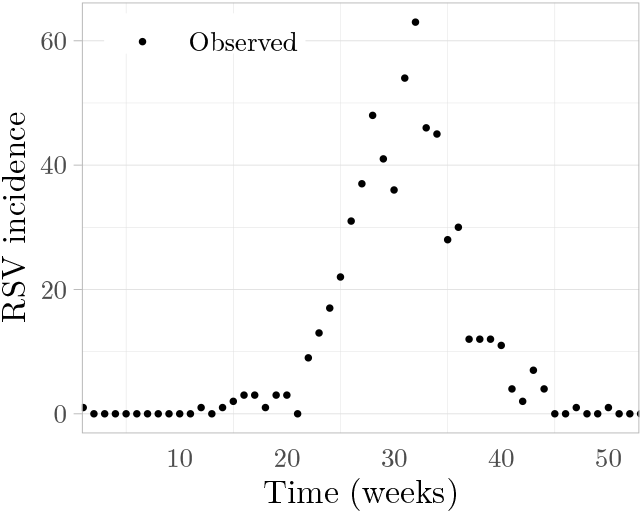
Incidence of RSV-associated hospitalizations.

To estimate the onset and offset for this RSV season, we begin by fitting a cyclic P-spline GAM (see Supplement) to the observations, using incidence at time *t, y*(*t*), as the model response variable, the log link function, and a 20-term cubic spline basis, *B*_*k*_ (*t*), as the only predictor such that

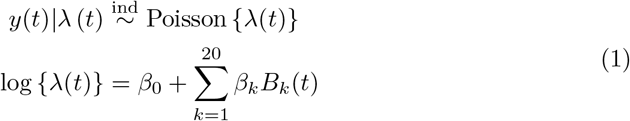

The best fit produced by this model (using a 2nd degree smoothing penalty) is shown in Fig 2a.

**Fig 2.**
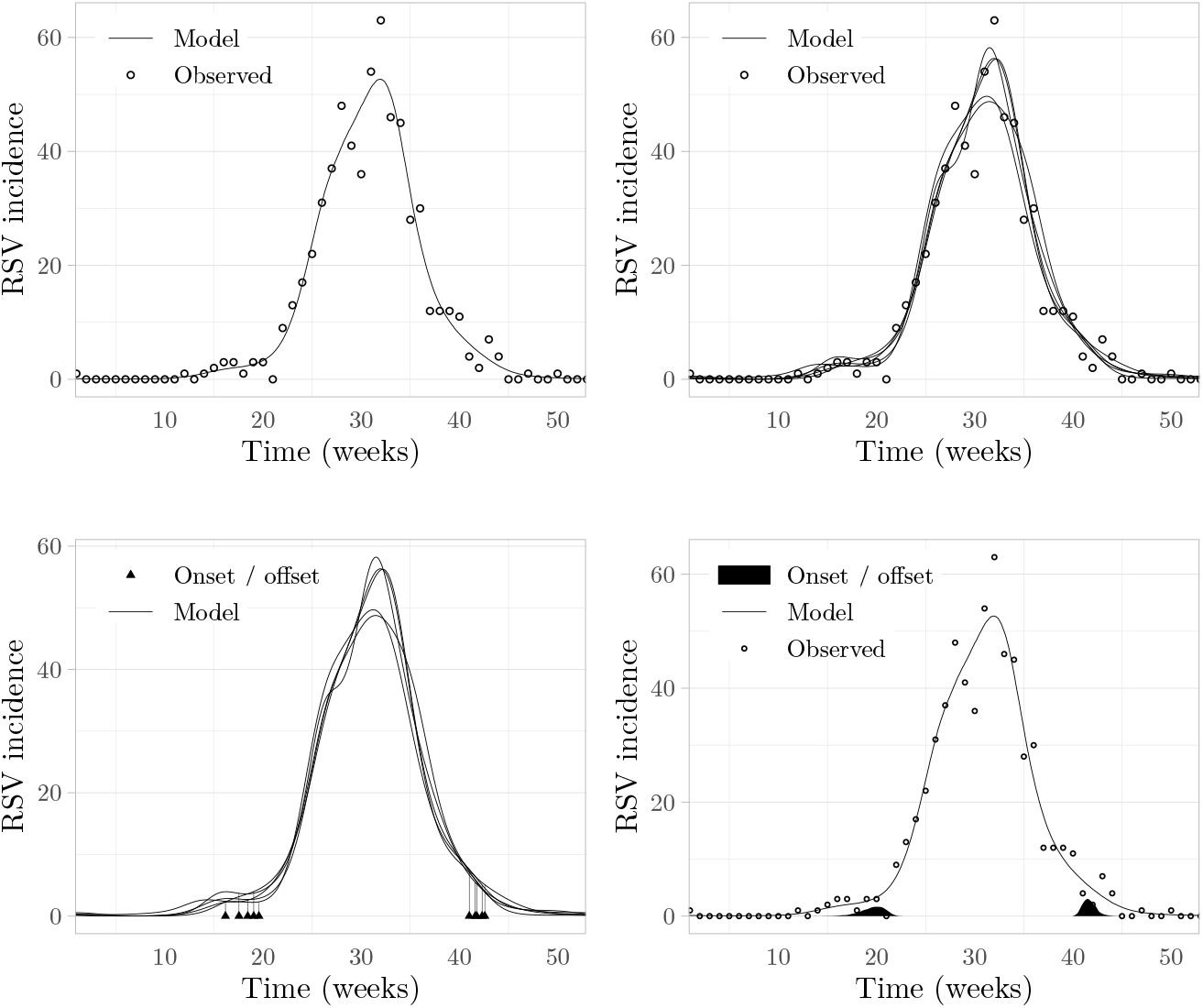
The steps involved in outcome estimation using penalized splines, demonstrated by estimating onset and offset of seasonal outbreaks of RSV. (a) Best-fit P-spline model of RSV incidence; (b) Five different sampled P-spline models of RSV incidence; (c) Five different computed estimates of RSV season onset and offset; (d) Sampled distribution of RSV season onset and offset estimates.

Using the asymptotic normality of the large-sample posterior distribution of *β*_*k*_, we can use parameter estimates (and uncertainty estimates) from the P-spline GAM to obtain a large sample credible interval for *λ*(*t*) by sampling *β*_*k*_ from this distribution and calculating 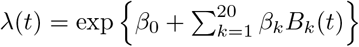 The resulting large sample credible interval has been shown to closely approximate the frequentist confidence interval across the entire function for non-linear data that follows a Poisson, binomial, or gamma distribution [10, 11]. Fig 2b demonstrates this in the form of *N* = 5 *λ*(*t*) curves produced by independently sampling each *β*_*k*_ *N* = 5 times; as *N* increases, the sampled curves yield large sample confidence intervals for *λ*(*t*) across all values of *t*.

Having obtained a smooth model with confidence intervals of the expected incidence, we can turn to our first outcome measures of interest, time of outbreak onset (*t*_*on*_) and offset (*t*_*off*_), defined by:

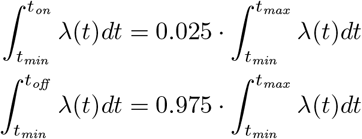

Given any *λ*(*t*) sampled via *β*_*k*_ sampling, we numerically solve for the corresponding *t*_*on*_ and *t*_*off*_, thereby indirectly sampling *t*_*on*_ and *t*_*off*_ from their sampling distributions. The same five *y*(*t*) with their corresponding *t*_*on*_ and *t*_*off*_ are shown in Fig 2c.

Sampling a large number of *t*_*on*_ and *t*_*off*_ yields an approximation of the unknown distributions of *t*_*on*_ and *t*_*off*_. In our example, sampling *N* = 2000 times yields the distributions of *t*_*on*_ and *t*_*off*_ shown in Fig 2d. From these distributions, we can calculate our desired point estimates (median) and interval estimates (quantiles): 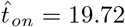 weeks (95% CI: 16.52 - 21.5 weeks), 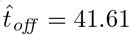 weeks (95% CI: 40.52 − 43.26 weeks).

We defined our other outcome measure, the preventable fraction (*F*_*p*_) of a prophylaxis regimen that offers protection between *t*_*start*_ and *t*_*end*_, as:

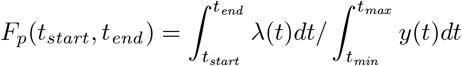

We repeated the above method to estimate the preventable fraction of the prophylaxis regimen recommended by the American Academy of Pediatrics as well as several alternative prophylaxis regimens, which then enabled us to assess the potential benefit of adjusting the prophylaxis regimen to better align with the local RSV season.

### Simulation study

We began by generating *N*_*season*_ = 60 RSV seasons. Each season was generated as follows:

1. Randomly choose epidemiologic week of season start *t*_*start*_ *∼* Uniform(5, 15), epidemiologic week of season end *t*_*end*_ *∼* Uniform(30, 45), and peak incidence *λ*_*max*_ *∼* Uniform(50, 500).
2. Calculate

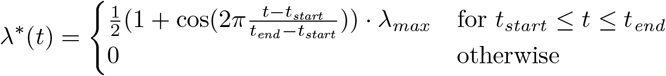

to produce a curve roughly of the desired shape (rising from 0 to *λ*_*max*_, then falling back to 0, between *t*_*start*_ and *t*_*end*_).
3. Fit a log-linked cubic P-spline to *λ*^*∗*^(*t*) to obtain true instantaneous incidence *λ*_0_(*t*); this ensures that the true incidence is smooth in the second derivative.

The number of seasons was chosen to balance power of the simulated outcome estimation with computational demand. The ranges of the parameters for the simulated seasons were chosen to include the observed characteristics in our actual dataset of RSV-related hospitalizations in Connecticut.

For each season, we then generated *N*_*obs*_ = 60 sets of observations by sampling observed incidence 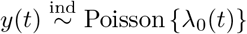 at weekly intervals. We also calculated the true values of *t*_*on*_ and *t*_*off*_ directly from *λ*_0_ (*t*) for each season.

Finally, we used the method described above to obtain 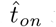 and 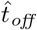 estimates for each of the *N*_*season*_ · *N*_*obs*_ = 3600 sets of observations, in order to verify that the true values of *t*_*on*_ and *t*_*off*_ calculated directly from *λ*_0_(*t*)) were contained in the 95% CI *≈* 95% of the time. We repeated the same process to validate the preventable fraction estimates.

## Results

To evaluate the results of our method, we considered three performance metrics:

- Coverage: the fraction of estimated 95% confidence intervals that include the true value of the outcome measure.
- Mean absolute error (MAE): the mean absolute value of the difference between point estimates and the true value.
- Scaled MAE (for onset and offset only): mean absolute error divided by *t*_*off*_ *− t*_*on*_.

The process of performing a simulation study is straightforward, but the measures of robustness it yields must be interpreted in the context of the research at hand. In our particular case, the MAE of onset/offset estimates (*∼* 0.15 weeks, see Table 1) was small compared to time periods of interest in our analysis (which were 1 weeks or more). The MAE for preventable fraction was similarly non-consequential to our results. Overall, the simulation study showed that our estimation method for onset, offset, and preventable fraction met the needs of our RSV research.

**Table 1.**
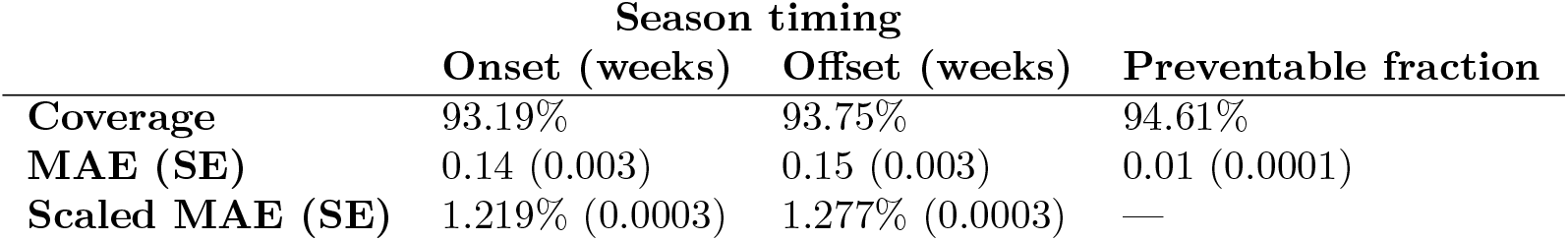
Simulation study results. MAE: Mean absolute Error. SE: Standard error.

## Discussion

In this study, we demonstrate how P-spline regression, combined with parameter resampling, can be used to generate uncertainty intervals for quantities derived from epidemic disease data. This is a general approach that can be applied to other to other systems, such as outbreaks of other infectious diseases, as well as other outcome measures.

We focus on specific outcome measures (onset/offset time and preventable fraction) that are relevant for RSV. Validating our estimation of those quantities using a simulation study gives methodological grounding to this approach. Direct public health applications of this approach include investigation of the potential benefits of region-specific RSV prophylaxis guidelines.

To facilitate application of P-spline estimation to other research, we wrote an R package, `pspline.inference` [12], that encapsulate our methods in an easy-to-use programming interface. It allows the user to obtain interval estimates of a user-defined outcome measure (as long as it’s computable from the model response variable), regardless of how complex its computation; it also provides an easy way to conduct a simulation study to validate estimation of the user-defined outcome measure on user-supplied data, thereby making it simple to develop and validate novel outcome measures. The package is available on GitHub and the Comprehensive R Archive Network (CRAN), and its documentation includes examples beyond those shown here.

We considered including a head-to-head comparison between our approach and any number of alternative approaches to estimation of time-based outbreak characteristics. However, such comparison would have required defining what it means to compare a point-estimation method to an interval-estimation method and what it means to compare two methods of estimating onset and offset in the absence of a standardized meaning of onset and offset. We therefore decided head-to-head comparison was not within the scope of this initial report, and instead we focus here on an absolute evaluation of the correctness of our approach by means of the simulation study.

The flexibility of our approach does come at a price. We validated it only for our input data and for our outcome measures; applications of this approach to other types of analysis will have to be validated independently. Our R package makes this validation simple to perform, but the validation is inherently time-consuming; whereas our analysis took minutes-to-hours to run, validation of our analysis took hours-to-days. Even with that computational cost, our approach is computationally less demanding than Bayesian Markov chain Monte Carlo (MCMC) approaches on datasets of comparable size, because the computationally expensive step in our approach is validation, not the analysis itself.

The estimates produced by our method are likely to be biased (as shown in our RSV application); measuring that bias (using our validation tools) and assessing its impact on results is, necessarily, left up to the individual application.

Our method is inherently retrospective; the strengths of our approach come from considering the incidence time series as a whole, and it therefore cannot, in its current form, be applied to real-time analysis.

In summary, ours is a method of retrospective analysis of infectious disease outbreaks based on P-splines; it is computationally less demanding than Bayesian MCMC methods, yet, unlike the common frequentist approaches, it allows for more straightforward interval estimation of time-based outbreak characteristics. Although we only validated it for our analysis of RSV seasonality, it is applicable to other similar systems, and by publishing it on CRAN, we hope to facilitate its use in other analyses.

## Data Availability

Data availability: not applicable
Code available at https://github.com/weinbergerlab/pspline.inference

## Supplement: P-spline GAMs in brief

A *spline* of order *n* is a piecewise polynomial real function *S*(*x*) of degree *n −*1 on interval [*x*_*L*_, *x*_*R*_]; the points *p*_1_, *p*_2_,…, *p*_*m*_ *∈* [*x*_*L*_, *x*_*R*_] at which its polynomial segments join are known as its *knots*. Owing to its construction, a spline of order *n* has up to *n −* 1 continuous derivatives throughout [*x*_*L*_, *x*_*R*_]; for our analysis, we only consider splines with exactly *n −* 1 continuous derivatives.

For any set of knots *P* = {*p*_1_,…, *p*_*m*_}, we can construct a unique set of *m − n* splines of order *n* known as the *basis splines (B-splines) over P*, denoted with *B*_*k*_(*x*; *n*; *P*), *k* = 1,…, *m − n*. Every spline *S*(*x*) of order *n* with knots *P* can be uniquely represented as a linear combination of the basis splines *B*_*k*_(*x*; *n*; *P*) such that:

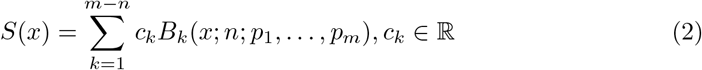

A *cyclic spline* of order *n* is a piecewise polynomial periodic real function *f* (*x*) = *f* (*x − T*). As with non-cyclic splines, a cyclic spline of order *n* is continuous in up to *n −* 1 derivatives, and we only consider those continuous in exactly *n −* 1 derivatives. Use of cyclic splines is often necessary in infectious disease modeling because non-cyclic splines do not guarantee smoothness at *x*_*L*_ *± kT* when extended to multiple periods. Basis splines of cyclic splines are themselves cyclic.

A *generalized additive model* provides a framework for modeling a response variable *y*, using predictor variables *x*_*i*_, *i* = 1,…, *p*, with corresponding potentially non-linear predictor functions *f*_*i*_, *i* = 1,…, *p*, and a link function *g*, such that the expected value, *E*(*y*), is defined as

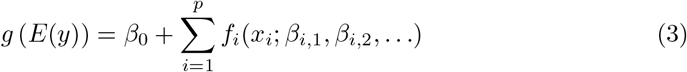

where *β*_0_ and *β*_*i,j*_ represent unknown model parameters.

Splines can be used for each predictor function in a GAM by setting:

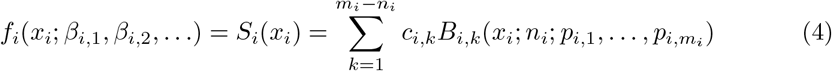

and treating the spline linear coefficients *c*_*i,k*_ as the model parameters *β*_*i,j*_. Splines for different *f*_*i*_ in the model can have different orders *n*_*i*_ and different knot sets *P*_*i*_ of length *m*_*i*_. After expanding each predictor spline, the GAM takes the form

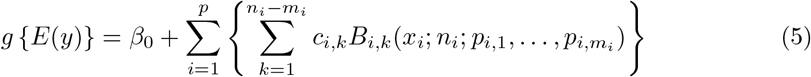

with *β*_0_ and *β*_*i,j*_ = *c*_*i,k*_ as model parameters, and *n*_*i*_ and *P*_*i*_ chosen prior to analysis.

This model can be fitted to data simply by minimizing ‖*y − E*(*y*) ‖ ^2^; however, this approach often leads to overfitting, especially with a larger number of knots. To mitigate this, the model can be modified to instead minimize ‖*y − E*(*y*) ‖ ^2^ + *P*, where *P* is a *penalty function* that measures non-smoothness of the model response variable. This approach balances closeness of fit with smoothness. B-splines used in a model with a penalty function are known as *penalized B-splines* (P-splines) [9]. A variety of penalty functions have been defined; the most common ones use *p*-th order differences between the model and the data, for some small integer *p* (often *p* = 2). As with general cyclic splines, *cyclic P-splines* are an extension of P-splines to periodic functions.

